# Diurnal rhythms in chimeric antigen receptor T cell performance: an observational study of 670 patients

**DOI:** 10.1101/2025.05.15.25327612

**Authors:** Patrick G Lyons, Emily Gill, Prisha Kumar, Melissa Beasly, Brenna Park-Egan, Zulfiqar Lokhandwala, Brandon Hayes-Lattin, Catherine L Hough, Nathan Singh, Guy Hazan, Huram Mok, Janice Huss, Colleen A McEvoy, Jeffrey Haspel

## Abstract

**Background:** Chimeric antigen receptor (CAR) T cells are a leading immunotherapy for refractory B-cell malignancies, but their impact is constrained by toxicity and incomplete long-term efficacy. Daily (circadian) rhythms in immune function may offer a lever to boost therapeutic success. Studies suggest that time of day influences immune-based therapies, including vaccines, hematopoietic stem cell transplantation, and checkpoint inhibitors for cancer. However, the clinical relevance of biological rhythms to CAR-T cell therapy remains unknown.

**Methods:** We retrospectively analyzed CAR-T cell survival and complications based on infusion time at two geographically distinct hospitals in St. Louis, Missouri (n=363), and Portland, Oregon (n=307) between 2018 and 2024. The primary outcome was 90-day overall survival (OS). Secondary outcomes included event-free survival (EFS), cytokine release syndrome (CRS), immune cell-associated neurotoxicity syndrome (ICANS), ICU admission, shock, respiratory failure, and infection. We quantified the independent relationship between infusion time and outcomes using multivariable mixed effects logistic regression, adjusting for patient, oncological, and treatment characteristics.

**Findings:** Morning CAR-T cell infusions were associated with higher overall survival and lower rates of neurotoxicity before and after adjusting for confounders. Each hour earlier in the day that CAR-T cell treatment was given corresponded to a 22% increase in the odds of 90-day OS (adjusted odds ratio [aOR] 0·78, 95% CI 0·66–0·92, p=0·004). Simultaneously, for every hour CAR-T cell infusion was delayed, the adjusted odds of severe ICANS rose by 17% (aOR 1·17, 95% CI 1·01–1·34, p=0·031), and anakinra prescription rose by 26% (aOR 1·26, 95% CI 1·07–1·49, p=0·006). In contrast, we did not find an association between infusion time and severe CRS (aOR 0·96, 95% CI 0·74–1·23, p=0·73). Temporal patterns were most pronounced in women and patients receiving CD19-targeting CAR-T cell products for leukemia or lymphoma.

**Interpretation:** Time of day is a potent and easily modifiable factor that could optimize CAR-T cell clinical performance.

**Funding:** National Institutes of Health.

**Research in context:** *Evidence before this study:* A review of PUBMED-cited articles shows that fundamental immune processes exhibit daily (circadian) rhythms in activity, leading immune-based therapies to vary in clinical efficacy based on the time of day. For example, the effectiveness of vaccines, checkpoint inhibitor therapy for cancer, and hematopoietic stem cell transplantation varies with the time of day they are administered to patients. One immunotherapy where dosing time remains unexplored in clinical data is CAR-T cells, a cell-based treatment for refractory B-cell cancers. Currently, the timing of CAR-T therapy is largely determined by staff availability. We hypothesized that CAR-T cell clinical safety and efficacy vary with administration time. If this hypothesis is correct, scheduling CAR-T cell infusions for specific times of day might improve the performance of this key treatment at minimal cost.

*Added value of this study:* This study reveals that the clinical performance of CAR-T cells varies substantially with administration time. It informs practice by showing that morning CAR-T cell infusions correlate with optimal therapeutic index based on better survival and less neurotoxicity. The study also identifies women and patients receiving anti-CD19 CAR-T cells as subgroups that may benefit the most from timed CAR-T cell infusions.

*Implications of all the available evidence:* This study on CAR-T cells should be understood in the context of growing evidence that circadian rhythms in immunity are broadly clinically translatable. The findings should prompt prospective trials that test time-of-day prioritization and, if validated, changes to CAR-T cell practice patterns. The findings also have implications for clinical trials applying CAR-T cells to new indications. Without controlling for time-of-treatment, diurnal rhythms in CAR-T cell effectiveness could be a significant confounder that could lead to false-negative or false-positive conclusions.

## INTRODUCTION

Chimeric antigen receptor (CAR) T cell therapies are becoming a mainstay treatment for refractory B-cell cancers and an emerging option in solid tumors and autoimmune diseases.^1^ CAR-T cells can deliver high response rates and durable remission; however, unmodifiable patient characteristics like age, performance status, and disease burden can mitigate CAR-T cell clinical effectiveness.^2^ Other determinants of effectiveness (e.g., product design, manufacturing, and induction chemotherapy regimen) require substantial effort and resources to alter. Beyond efficacy, these factors affect the risk of life-threatening complications of CAR-T cell therapy, including cytokine release syndrome (CRS) and immune effector cell-associated neurotoxicity syndrome (ICANS).^3^ To improve the clinical utility of CAR-T cells, we must identify easily modifiable factors that can optimize their therapeutic index.

One candidate factor is the time of day, which reflects circadian variation in the patient’s immune system.^4^ Circadian rhythms are 24-hour biological cycles generated by a conserved transcription factor network called the molecular circadian “clock.” Clinical evidence and pre-clinical animal models indicate that the clock imposes daily rhythms on fundamental immune processes, leading many immune-based therapies to vary in efficacy based on the time of day. For example, the effectiveness of the SARS-CoV-2 and varicella zoster virus vaccines differs with the time of immunization.^5,6^ Immune checkpoint inhibitors for cancer and hematopoietic stem cell transplants (HSCT) also perform differently based on infusion time.^7–10^ A preclinical study comparing CAR-T cell efficacy based on time-of-treatment found that CAR-T cells administered to mice at the end of their activity period (corresponding to evening in humans) failed to inhibit tumor growth.^11^

However, the translatability of these observations to real-world clinical practice remains unknown in terms of both CAR-T cell efficacy and toxicity. We therefore studied the relationship between the time of CAR-T cell infusion and patient outcomes, hypothesizing that earlier administration times would correlate with better clinical outcomes and fewer or less severe complications.

## METHODS

### Study Design, Setting, and Patients

This is a retrospective cohort study of all adult patients receiving a single treatment of CAR-T cells as a standard-of-care therapy for a hematologic malignancy at two NCI-designated Comprehensive Cancer Centers: Siteman Cancer Center at Washington University School of Medicine (WU, St. Louis, MO), and Knight Cancer Institute at Oregon Health & Science University (OHSU, Portland, OR), between January 1, 2018 and December 31, 2024. Relevant to circadian rhythms, which are sensitive to light-dark cycles, the two centers are situated at different latitudes (38·6°N for WU, 45·5°N for OHSU).

Indications for CAR-T cell treatment included diffuse large B-cell lymphoma, mantle cell lymphoma, follicular lymphoma, acute lymphocytic B-cell leukemia, and multiple myeloma. There were six FDA-approved CAR-T cell products used during the study period: axicabtagene ciloleucel (axi-cel), lisocabtagene maraleucel (liso-cel), tisagenlecleucel (tisa-cel), brexucabtagene autoleucel (brexu-cel), ciltacabtagene autoleucel (cilta-cel), and idecabtagene vicleucel (ide-cel).

During the study period, WU exclusively administered CAR-T cell therapy in the inpatient setting, while OHSU performed some infusions in the outpatient setting for selected patients.^12^ At both institutions, the timing of CAR-T cell administration primarily depends on the pharmacy logistics and clinical staffing (e.g., attending physician availability). We excluded patients who received multiple CAR-T cell infusions (**eFigure 1**). All patients had at least 90 days of follow-up after treatment.

The institutional review boards at Washington University School of Medicine (#202410178) and OHSU (#00027601) approved this study. Since this was a retrospective study, patient consent was not required. See the **appendix** for STROBE reporting documentation.

### Data

We extracted electronic health record (EHR; Epic, Verona, WI) data from each hospital’s research data warehouse. These included demographic and encounter data, vital signs, laboratory and microbiology tests, and medications. The primary outcome was 90-day overall survival (OS). Secondary outcomes were: 90-day event-free survival (EFS, defined based on relapse, progression, or death from any cause), 365-day OS and EFS (among patients with at least one year of observation time), the occurrence and timing of disease progression or relapse, development (and grade) of CRS or ICANS (based on American Society for Transplantation and Cellular Therapy consensus grading), new infection (defined as newly positive blood, respiratory, or urine culture, newly positive respiratory viral test, or newly positive test for *Clostridium difficile* infection within 14 days of CAR-T cell administration), intensive care unit (ICU) admission, invasive mechanical ventilation (IMV), and shock (receipt of vasoactive medications).^13,14^ In calculating survival measures from the time of CAR-T cell infusion, we censored patients who were alive and event-free at last follow-up or on April 1, 2025 (whichever came first).

The primary exposure was time of day, modeled as a continuous variable. To facilitate categorical comparisons in time-to-event analyses, we dichotomized time at 15:00. We chose this threshold based on practical relevance, as 15:00 approximates the midpoint of afternoon and early evening clinical activities (12:00–18:00). This time is also consistent with studies of immune checkpoint inhibitor and HSCT effectiveness as a function of infusion times.^7–9^ *A priori*, we analyzed infusion times based on conventional clock time and as hours after local sunrise (to adjust for latitude differences between the centers, seasonal changes to photoperiod, and Daylight Savings Time).^15^ This sensitivity analysis addresses biologic plausibility since circadian rhythms are strongly synchronized by sunlight.

### Analysis

We present data as n (%) and median (interquartile range [IQR]). We used Chi-square and Wilcoxon Rank-Sum tests for unadjusted comparisons, as appropriate.

In the primary analysis of OS and EFS, we used multivariable mixed-effects logistic regression models (with hospital as a random effect) to determine the adjusted relationship between time of day and dichotomous outcomes; each model yielded an adjusted odds ratio corresponding to each additional hour later in CAR-T cell administration. To enhance clinical interpretability of the landmarked survival models, we calculated the absolute risk reduction (ARR) at each hour (relative to 08:00), summarizing the mean ARR over this time window and obtaining confidence intervals via nonparametric bootstrap.

All multivariable models were adjusted for covariates selected *a priori* based on a directed acyclic graph informed by the literature, biological plausibility, and our clinical experience (**eTable 1**).^16^ These included age, sex, diagnosis indicating CAR-T therapy, lymphodepletion regimen, Eastern Cooperative Oncology Group (ECOG) performance status, comorbidity burden (van Walraven-weighted Elixhauser index), severity of illness on day-0 (Simplified Acute Physiology Score [SAPS]-2), CAR-T cell dose, temporal trends (season and calendar year within study), and time in hospital before infusion (defaulting to 0 days for outpatient administrations).^17–19^

Concurrently, we used Kaplan-Meier methods to estimate survival distributions for early and late CAR-T cell administrations and compared them using log-rank tests. To quantify absolute survival differences over a fixed time horizon, we compared each group’s 2-year restricted mean survival time (RMST).^20^ We then fit multivariable survival models to account for possible confounding. We initially used multivariable Cox models with hospital-clustered robust standard errors; however, Schoenfeld residuals indicated proportional hazards assumption violations (p<0·01). We therefore estimated covariate-adjusted 2-year RMST via the pseudo-observation approach; this approach yields an exposure regression coefficient that directly represents the adjusted difference in 2-year RMST per hour change in CAR-T cell administration time.^21^ For these models, we obtained confidence intervals via parametric bootstrap to account for model uncertainty and the small number of hospital clusters.

For each model in the main analysis, we calculated E-values to quantify the robustness of effect estimates to hypothetical unmeasured confounders.^22^ The E-value is an odds ratio representing the minimum strength of association that an unmeasured confounder would need to have (with both the exposure and the outcome) to negate the observed association.

We prespecified four subgroup analyses. First, we compared the findings between hospitals. Second, at OHSU, we compared inpatient versus outpatient CAR-T cell administrations; the latter were prescheduled and predominantly for low-risk patients, making it less likely that acute patient factors would influence time-outcome relationships.^12^ Third, since circadian patterns are sex-dimorphic, we stratified our analyses according to this variable.^23^ Fourth, we compared results for patients receiving B-cell maturation antigen (BCMA)-directed versus CD-19-directed CAR-T cells, hypothesizing that tumor-or treatment-specific factors may modify temporal effects.

We analyzed patterns in commonly used biomarkers based on the timing of CAR-T cell administration. We compared day-0, day-7, day-10, and day-14 absolute lymphocyte counts (ALCs), as well as peak values (within 14 days) of serum C-reactive protein (CRP), lactate dehydrogenase (LDH), and ferritin measurements between early (before 15:00) and late administration times. At OHSU, where additional biomarkers were routinely available from clinical care, we made analogous comparisons of IFN-γ, TNF-α, IL-1, IL-2, IL-5, IL-6, IL-7, IL-10, IL-12, and IL-13.

We regarded p-values <0·05 as statistically significant. All analyses used R 4·4·1 (R Project for Statistical Computing) and the following packages: tidytable, collapse, gtsummary, ggplot2, pseudo, lme4, and EValue.

## RESULTS

This study analyzed all patients receiving a single dose of CAR-T cells as standard-of-care treatment between January 1, 2018, and December 31, 2024, at two geographically distinct quaternary centers (**Table 1**). The cohort comprised 670 patients: 363 from Washington University in St. Louis, MO (WU), and 307 from Oregon Health & Science University (OHSU) in Portland, OR. Patient characteristics were similar between sites. However, there were differences between the centers in the distribution of malignancies, CAR-T cell products, and treatment setting (OHSU provided outpatient CAR-T cell infusions to a subset of patients, while infusions at WU were all inpatient). Both sites administered CAR-T cell infusions over an approximately eleven-hour span (range: 08:46-17:42, **Figure 1A**), with infusions occurring slightly earlier at WU than at OHSU (median 10:57 [IQR 10:21-12:03] vs 12:22 pm [11:41-13:40]). Despite these case-mix and practice differences, the two locations had similar overall and event-free survival at 90 and 365 days.

**Figure 1:**
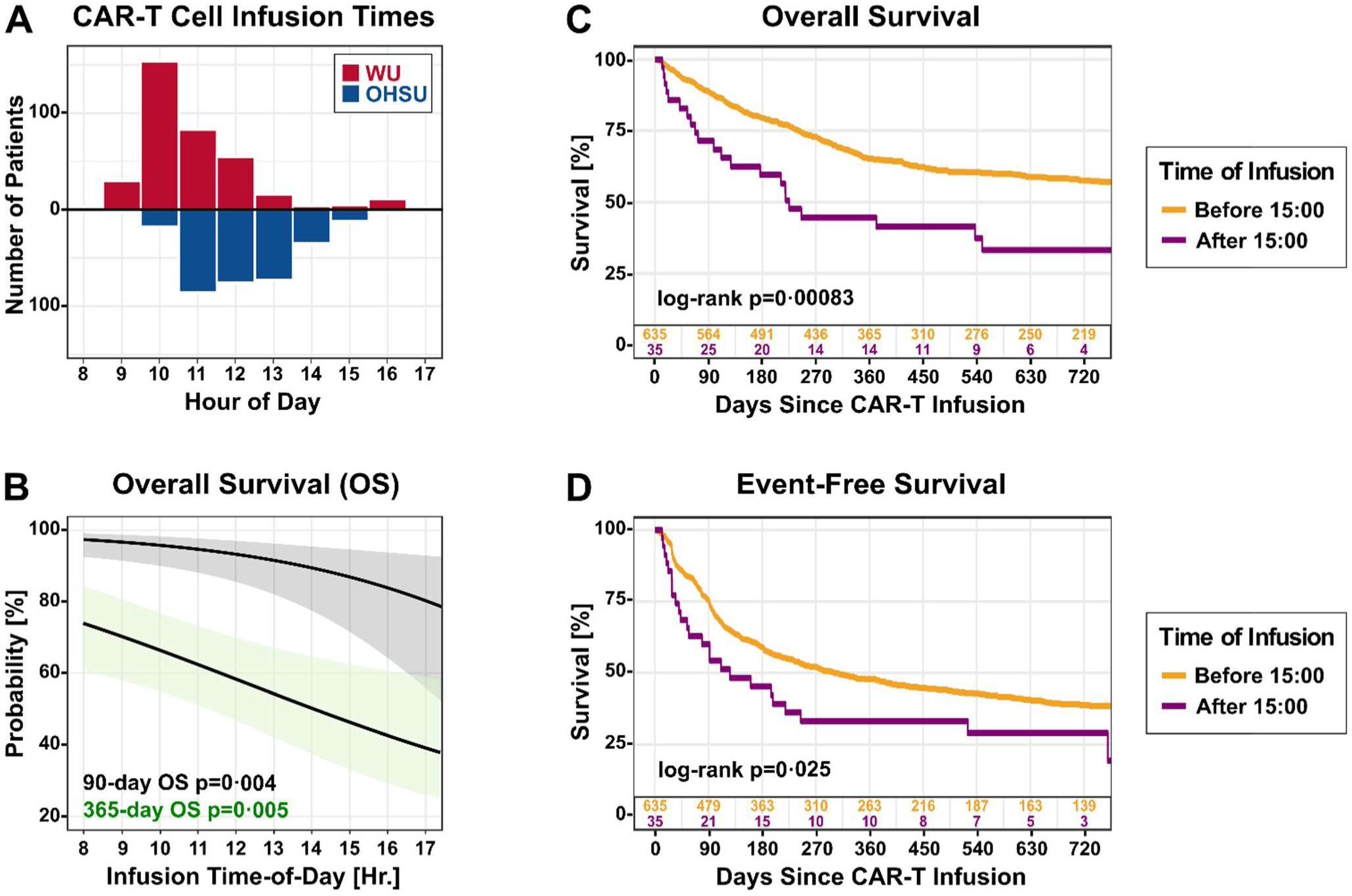
CAR-T cell treatment times vary and are associated with survival. **A** shows the distribution of CAR-T cell infusion times at WU (red bars) and OHSU (blue bars). **B** shows a multivariable mixed-effects logistic regression analysis relating CAR-T infusion time to 90-day OS (black line) and 365-day OS (green shaded line). Shaded areas represent 95%-CIs. The Wald test p-values are depicted. For depiction of EFS, see **eFigure 3**. **C** and **D** depict Kaplan-Meier analyses comparing CAR-T cell infusion times before 15:00 (orange lines, n=635) to after 15:00 (purple lines, n=35). **Panel C**, overall survival. **Panel D**, event-free survival.

**Table 1.** Characteristics of patients receiving CAR-T cell infusions at WU and OHSU.

| Characteristic | WU N = 363 | OHSU N = 307 |
| --- | --- | --- |
| Age, median (IQR) | 66 (58, 73) | 64 (55, 71) |
| Female, n (%) | 114 (31%) | 108 (35%) |
| Malignancy, n (%) |  |  |
| DLBCL | 236 (65%) | 174 (57%) |
| Other Lymphomas | 32 (8.8%) | 47 (15%) |
| Leukemia | 15 (4.1%) | 20 (6.5%) |
| Multiple Myeloma | 80 (22%) | 66 (21%) |
| Product |  |  |
| axi-cel | 186 (51%) | 64 (21%) |
| brexu-cel | 33 (9.1%) | 40 (13%) |
| cilta-cel | 47 (13%) | 29 (9.4%) |
| ide-cel | 33 (9.1%) | 37 (12%) |
| liso-cel | 29 (8.0%) | 50 (16%) |
| tisa-cel | 35 (9.6%) | 87 (28%) |
| Time of Day, median (IQR) | 10:57 (10:21, 12:03) | 12:22 (11:41, 13:40) |
| Hours After Sunrise, median (IQR) | 4.6 (3.6, 5.6) | 5.9 (4.9, 6.9) |
| Fludarabine/Cyclophosphamide Conditioning, n (%) | 329 (91%) | 226 (74%) |
| Outpatient CAR-T cell Infusion, n (%) | 0 (0%) | 65 (21%) |
| Hospital Days Before Infusion, median (IQR) | 0.9 (0.7, 3.0) | 0.8 (0.7, 0.9) |
| ECOG Performance Status, median, IQR) | 1 (0, 1) | 1 (1, 1) |
| Van Walraven Score, median (IQR) | 9 (9, 14) | 12 (2, 18) |
| SAPS-2, median (IQR) | 28 (28, 32) | 27 (24, 34) |
| Period, n (%) |  |  |
| 2018-2019 | 64 (18%) | 35 (11%) |
| 2020-2022 | 163 (45%) | 118 (38%) |
| 2023-2024 | 136 (37%) | 154 (50%) |
| Season, n (%) |  |  |
| Fall | 83 (23%) | 79 (26%) |
| Spring | 104 (29%) | 77 (25%) |
| Summer | 84 (23%) | 89 (29%) |
| Winter | 92 (25%) | 62 (20%) |
| **Patient Outcomes** |  |  |
| Overall survival, 90 days | 317 (87%) | 272 (89%) |
| Event-free survival, 90 days | 264 (73%) | 236 (77%) |
| Overall survival, 365 days* | 218 (63%) | 155 (62%) |
| Event-free survival, 365 days* | 152 (44%) | 116 (46%) |
| ICU admission, n (%) | 61 (17%) | 35 (11%) |
| Mechanical ventilation, n (%) | 28 (7·7%) | 10 (3·3%) |
| Vasopressors, n (%) | 45 (12%) | 20 (6·5%) |
| Infections, n (%) | 85 (23%) | 26 (8·5%) |
| CRS, n (%) |  |  |
| Any CRS | 297 (82%) | 237 (77%) |
| Grade 3/4 CRS | 29 (8·0%) | 14 (4·6%) |
| ICANS, n (%) |  |  |
| Any ICANS | 167 (46%) | 117 (38%) |
| Grade 3/4 ICANS | 72 (20%) | 52 (17%) |
| Tocilizumab, n (%) | 321 (64%) | 182 (14%) |
| Anakinra, n (%) | 29 (8·0%) | 44 (14%) |

\*WU N = 343, OHSU N = 250
IQR, interquartile range; DLBCL, diffuse large B-cell lymphoma; CAR T-cell, chimeric antigen receptor; ECOG, Eastern Cooperative Oncology Group; ICU, intensive care unit; CRS, cytokine release syndrome; ICANS, immune cell-associated neurotoxicity syndrome

We first examined the independent relationship between CAR-T cell outcomes and time of day through mixed-effects multivariable logistic regression, treating infusion time as a continuous variable and adjusting for key patient, oncologic, and treatment characteristics (**Figure 1B**). This model demonstrated an inverse relationship between the time of day CAR-T cells were infused and subsequent 90-day and 365-day overall survival (OS). Expressing infusion time relative to local sunrise rather than clock time (to adjust for differences in latitude between the centers, seasonal variation in photoperiod, and Daylight Savings Time) produced comparable results (**eFigure 2**). Event-free survival (EFS) also showed an inverse relationship with CAR-T infusion time in the model (**eFigure 3**).

To orthogonally validate these findings, we performed time-to-event analyses comparing survival in patients receiving CAR-T cell infusions before 15:00 (early, n=635) and after 15:00 (late, n=35). We chose 15:00 as a cutoff based on practical relevance (midpoint of afternoon clinic schedule) and consistency with studies of diurnal rhythms in checkpoint inhibitors.^7,9^ Patients were similar regardless of CAR-T cell administration time (**eTable 2**). Still, later infusions were associated with worse OS and EFS in Kaplan-Meier analysis (**Figures 1C and 1D**) and survival landmarked at 90 days (n=25/35 [71%] vs n=564/635 [89%], p=0.006) or 365 days (n=14/33 [42%] vs n=364/565 [64%], p=0.011) (**Table 2**).

**Table 2.** Outcomes of patients receiving CAR-T cell infusions before and after 15:00.

| **Outcome** | **08:00-14:59, N = 635** | **15:00-18:00, N = 35** | **p-value** |
| --- | --- | --- | --- |
| ***Survival Outcomes*** |  |  |  |
| 90-Day Overall Survival, n (%) | 564 (89%) | 25 (71%) | 0·006 |
| 365-Day Overall Survival,<sup>a</sup> n (%) | 364 (64%) | 14 (42%) | 0·011 |
| 90-Day Event-Free Survival, n (%) | 479 (75%) | 21 (60%) | 0·041 |
| 365-Day Event-Free Survival,<sup>a</sup> n (%) | 263 (47%) | 10 (30%) | 0·069 |
| 2-year RMST, days, median (IQR) | 515 (493, 537) | 358 (259,458) | 0·003 |
| ***Complications*** |  |  |  |
| ICU Admission, n (%) | 86 (14%) | 10 (29%) | 0·013 |
| Mechanical Ventilation, n (%) | 31 (4·9%) | 7 (20%) | 0·002 |
| Vasopressors, n (%) | 57 (9·0%) | 8 (23%) | 0·014 |
| CRS, n (%) | 506 (80%) | 28 (80%) | >0·9 |
| Grade 3/4 CRS, n (%) | 37 (5·8%) | 6 (17%) | 0·019 |
| ICANS, n (%) | 265 (42%) | 19 (54%) | 0·14 |
| Grade 3/4 ICANS, n (%) | 113 (18%) | 11 (31%) | 0·043 |
| Any Infection, n (%) | 98 (15%) | 13 (37%) | <0·001 |
| Bloodstream Infection, n (%) | 42 (6·6%) | 4 (11%) | 0·3 |
| Respiratory Infection, n (%) | 37 (5·8%) | 8 (23%) | 0·001 |
| Genitourinary Infection, n (%) | 7 (1·1%) | 0 (0%) | >0·9 |
| C difficile, n (%) | 21 (3·3%) | 3 (8·6%) | 0·12 |
| <sup>a</sup>365 days of observation time available for 565 and 33 patients, respectively |  |  |  |
| CAR T-cell, chimeric antigen receptor T-cell; RMST, restricted mean survival time; ICU, intensive care unit; CRS, cytokine release syndrome; ICANS, immune effector cell-associated neurotoxicity syndrome |  |  |  |

To better understand the practical implications of our data, we investigated how a one-hour shift in CAR-T cell infusion time relates to outcomes (**Figure 2A, eTables 3 and 4**). In our multivariable regression model, every hour earlier that CAR-T cell treatment was given, the adjusted odds ratio of 90-day survival rose by 21% (aOR 0.78, 95% CI 0·66–0·92, p=0·004), and 365-day survival rose by 16% (aOR 0·84, 95% CI 0·74– 0·95, p=0·005). For our cohort, this translates into a 7.1% average absolute reduction in 90-day survival (95% CI 3·5-11·1%) for each hour that CAR-T cell infusion is delayed. The corresponding E-values for these estimates (i.e., the minimum strength of unmeasured confounding needed to account for the observed association, expressed as an odds ratio) were 1·50 for 90-day OS and 1·42 for 365-day OS. Given that these values reflect a small, one-hour time difference, substantial unmeasured confounding would be needed to dismiss our observations. In adjusted time-to-event models (**eTable 3**), each hour later in infusion time was associated with a 22-day reduction in 2-year overall restricted mean survival time (RMST, 95% CI 8·1 to 36-day reduction) and a similar decrease in event-free RMST (18-day decrease per hour, 95% CI 2·2 to 34-day reduction). These values represent absolute differences of approximately 3% per hour, a magnitude resembling that found in logistic regression analyses. Altogether, even relatively small changes in CAR-T infusion times are associated with clinically meaningful differences in survival.

**Figure 2:**
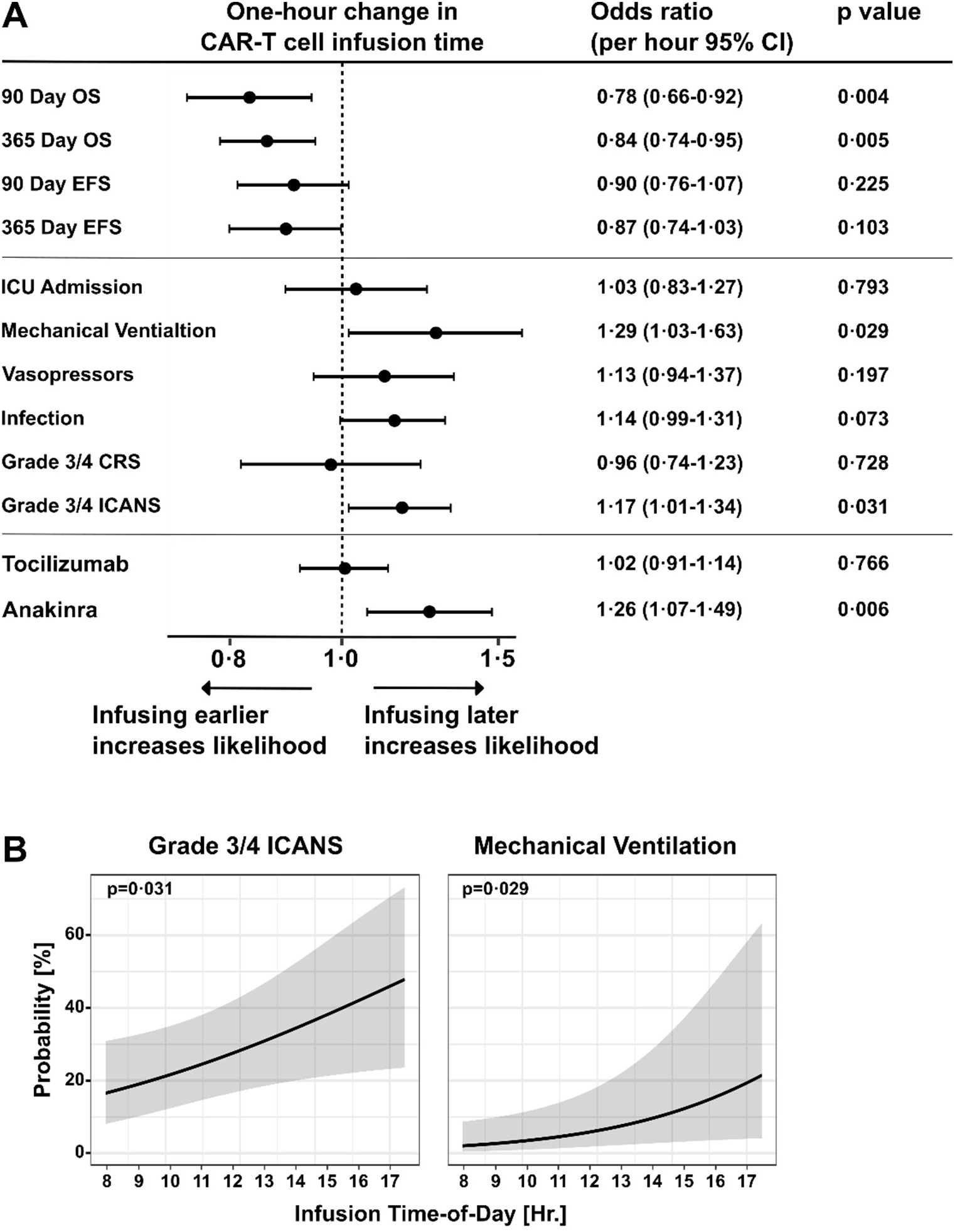
A one-hour shift in CAR-T cell infusion time significantly correlates with efficacy and safety. **A** depicts hourly adjusted odds ratios ± 95%-CIs estimated by multivariable mixed-effects logistic regression for the indicated outcomes. The area left of the dashed line signifies that an event would be more likely to happen if CAR-T cell infusions were shifted one hour earlier; to the right of the dashed line indicates increased odds of the event happening if CAR-T cell infusions were shifted one hour later. **B** shows a multivariable mixed effects logistic regression analysis relating CAR-T infusion time to the probability of severe (grade 3/4) ICANS (left panel) or mechanical ventilation (right panel). Shaded areas represent 95%-CIs. The Wald test p-values are depicted. For modeling of other adverse outcomes, see **eFigure 4.**

In addition to survival, adverse events also correlated with the timing of CAR-T cell infusions. In the multivariable regression analysis these included severe neurotoxicity (**Figures 2A and 2B, left panel**), as measured by the incidence of grade 3-4 ICANS (hourly aOR 1·16, 95% CI 1·01–1·33, p=0·042), and anakinra prescription (hourly aOR 1·26, 95% CI 1·06–1·49, p=0·008). The likelihood of invasive mechanical ventilation after CAR-T cell treatment also increased as infusion times grew later in the day (**Figure 2B, right panel**). In contrast, we did not find an association between infusion time and severe CRS (hourly aOR 0·96, 95% CI 0·76–1·22, p=0·74, **eFigure 4 and eTable 4**). Time-to-event analyses yielded similar results, comparing adverse events in patients receiving CAR-T cell infusions before and after 15:00 (**Table 2**). Late CAR-T infusion correlated with increased rates of mechanical ventilation (n=7 [20%] vs n=31 [4·9%], p=0·002), vasopressor use (n=8 [23%] vs n=57 [9·0%], p=0·014), infection (n=13 [37%] vs n=98 [15%], p<0·001), severe (grade 3/4) CRS (n=6 [17%] vs n=37 [5·8%], p=0·019), and severe ICANS (n=11 [31%] vs n=113 [18%], p=0·043). Altogether, our results indicate that CAR-T cell infusions in the morning had the best therapeutic index based on better survival and lower rates of severe neurotoxicity. Nevertheless, shifting infusions earlier at any point in the day was associated with improved performance.

Clinicians typically measure blood cell counts and serologic markers of inflammation after CAR-T cell treatments.^24,25^ As such, we investigated whether standard biomarkers would be sensitive to CAR-T cell infusion time. The recovery rate in absolute lymphocyte count, a predictor of CAR T efficacy, did not differ based on infusion time through day 14 **(eFigure 5)**.^25^ Inflammatory serologies – CRP, LDH, and ferritin – were available for most patients (n=661), and a subset of inpatients in the OHSU cohort had measurements of circulating cytokines (n=167). While we observed slightly increased maximal ferritin and IL-6 levels with late CAR-T infusions (after 15:00), these were not statistically significant (**eTable 5**).

Finally, we performed pre-specified subgroup analyses of treatment site (WU vs OHSU), treatment setting (inpatient vs outpatient), sex, and CAR-T cell target. The relationship between CAR-T infusion time and survival was similar across treatment sites and settings (**eFigure 6**). In contrast, sex-based subgroup analysis suggested a stronger association between CAR-T cell timing and outcomes in female patients (**Figure 3**). In our multivariate logistic regression model, the slope of the relationship between adjusted 90-day OS and CAR-T cell infusion time was steeper for females than males (**Figure 3A**). On a per-hour basis, the odds of 90-day OS were stronger among female CAR-T cell recipients (aOR 0·57, 95% CI 0·39-0·85) than among male recipients (0·86, 95% CI 0·70-1·05), although the interaction term was not statistically significant (p=0·32). Comparing early (before 15:00) versus late (after 15:00) infusions via Kaplan-Meier analysis further supports the conclusion that female patients derive more benefit than males from earlier CAR-T cell administration times (**Figure 3B**). Separately, subgroup analysis based on CAR-T cell target suggested differential time-sensitive patterns between CD-19- and BCMA-directed therapies (**Figure 4**). Patients receiving anti-CD19 CAR-T cells (for leukemia and lymphoma) exhibited a strong time-outcome relationship, while anti-BCMA CAR-T cells (for multiple myeloma) appeared less time sensitive.

**Figure 3:**
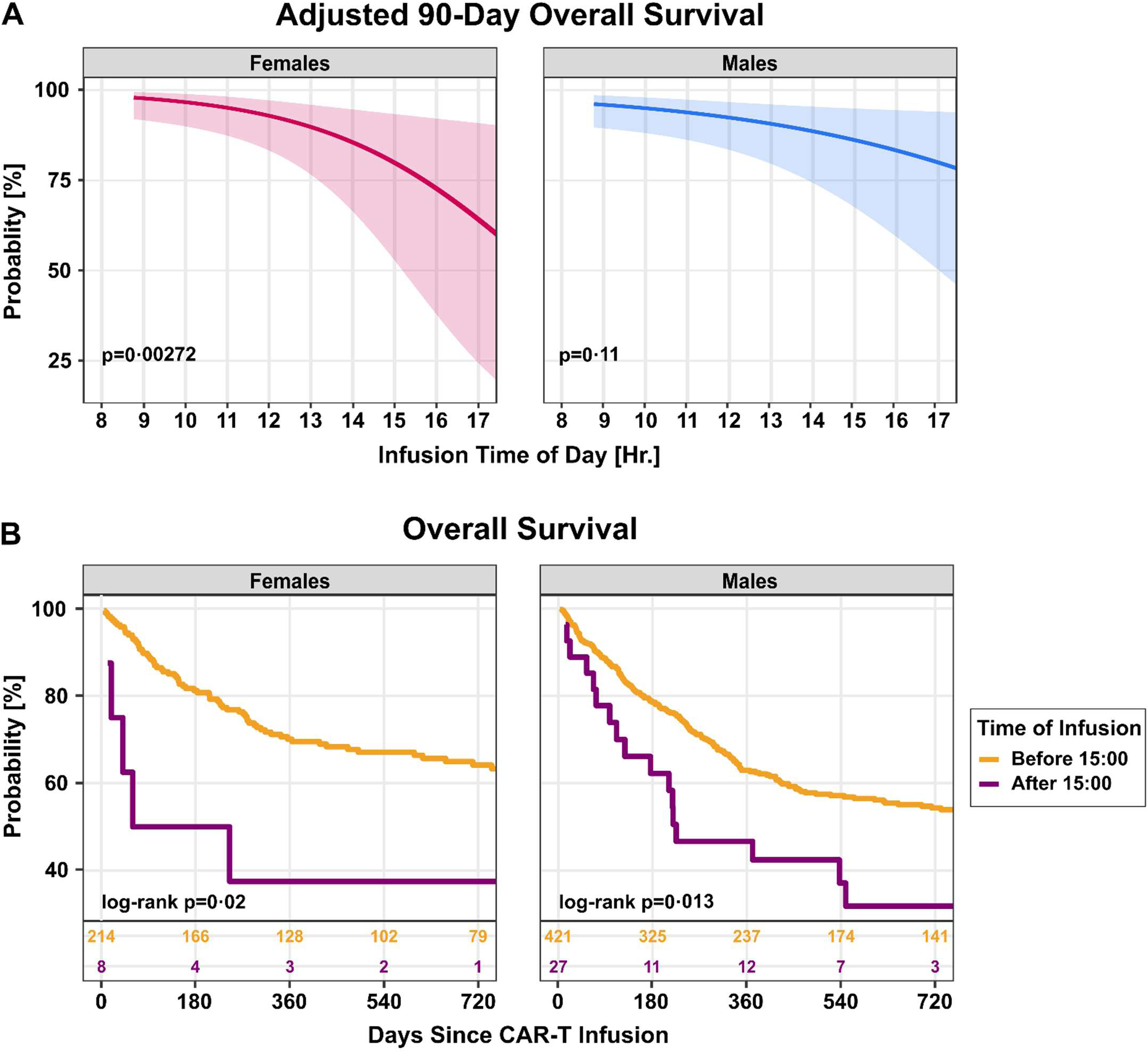
CAR-T cell outcomes are more time-sensitive in female patients. **A** is a multivariable mixed-effects logistic regression analysis relating CAR-T infusion time to 90-day survival for female patients (n=222, left panel, pink line), and male patients (n=448, right panel, blue line). Shaded areas represent 95%-CIs. Wald test p-values for trend are depicted. **B** depicts a Kaplan-Meier analysis of overall survival binned by infusion time of day and sex. Left panel, female patients (n=222). Right panel, male patients (n=448). Orange, infusions are given before 15:00. Purple, infusions given after 15:00. Log-rank p values and the number of patients at risk are provided within each graph.

**Figure 4:**
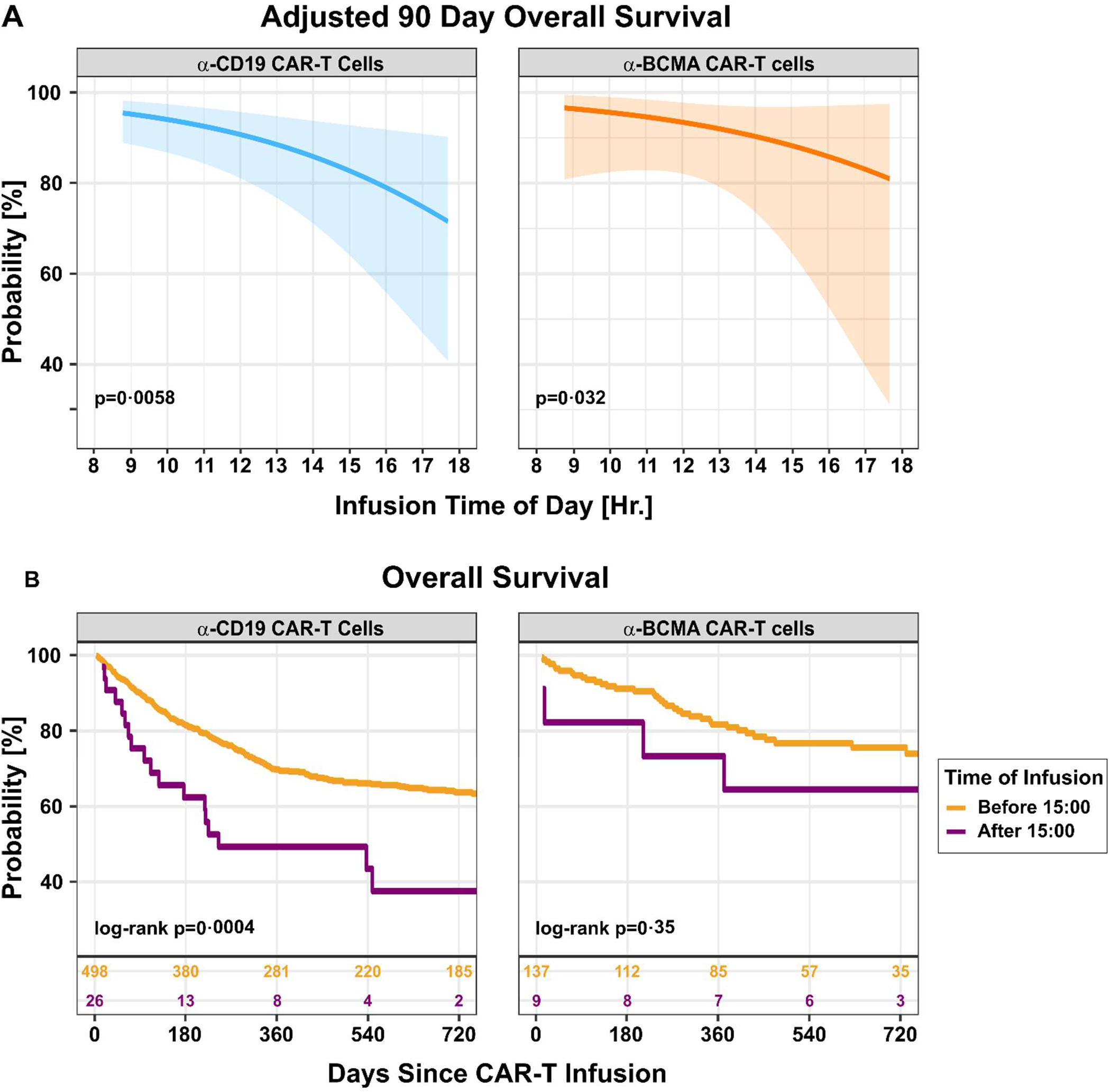
CAR-T cell outcomes are more time-sensitive in patients receiving anti-CD19 T cells. **A** is a multivariable mixed-effects logistic regression analysis relating CAR-T infusion time to 90-day survival for patients receiving anti-CD19 CAR-T cells (left panel, n=524) and patients receiving anti-BCMA cells (right panel, n=146). Shaded areas represent 95%-CIs. Wald test p-values for trend are depicted. **B** depicts a Kaplan-Meier analysis of overall survival binned by infusion time of day and CAR-T cell antigen specificity. Left panel, patients receiving anti-CD19 CAR-T cells (n=524). Right panel, patients receiving anti-BCMA cells (n=146). Orange, infusions are given before 15:00. Purple, infusions are given after 15:00. Log-rank p values and the number of patients at risk are depicted within each graph.

## DISCUSSION

In this multicenter study with real-world patient data, we observed a strong association between CAR-T cell infusion timing and clinical outcomes, with earlier infusions having better survival and lower rates of severe neurotoxicity. These associations were reproducible across treatment centers and were robust to multiple sensitivity analyses. Our results should be understood in the context of growing evidence that circadian rhythms in immunity are broadly clinically translatable. Clinical and preclinical studies of vaccination, checkpoint inhibitor cancer therapy, and HSCT point to a conserved benefit in scheduling immunotherapy to coincide with the circadian onset of wakefulness (mornings in humans, evenings in mice).^5–11^ Our study suggests that biological rhythms extend to CAR-T cell clinical performance.

The clinical impact of our findings may be substantial for patients and health systems if treatment time proves causal. Each hour delay in CAR-T cell infusion corresponded to a 7% average decrease in 90-day overall survival in our adjusted model (i.e., one-hour earlier infusions for 14 patients would prevent one additional death)– a magnitude on par with the survival reductions observed with delayed antibiotics in septic shock and delayed revascularization in ST-elevation myocardial infarction.^26,27^ In those scenarios, such patterns prompted major clinical practice changes aimed at minimizing time to treatment. With these examples as precedent, our findings should prompt prospective trials evaluating time-of-day prioritization strategies, with an eye towards modifying CAR-T cell therapy practice patterns if validated.

Our results also have implications for clinical trial design, which is important for evaluating new CAR-T cell products and new indications like solid tumors, autoimmune disorders, and interstitial lung disease. Without controlling for time-of-treatment, diurnal rhythms in CAR-T cell effectiveness could be a significant confounder that leads to false-negative or false-positive conclusions. This mechanism has been postulated for the translational failure of some novel stroke treatments.^28^ Indeed, because trial protocols often require additional pre-infusion procedures, coordination, or regulatory steps, investigational CAR-T cell treatments may be more likely to occur later in the day than their standard-of-care counterparts, potentially obscuring the efficacy of novel therapies.

One potential challenge to implementing CAR-T cell chronotherapy involves operational inefficiencies caused by compressing treatments into a shorter interval of the day. However, these inefficiencies might be offset by improvements in patient outcomes and reductions in costly interventions such as rescue therapies like anakinra. Further, subgroup analysis suggests that selectively targeting women and patients with anti-CD19 treatments (lymphoma and leukemias) for early infusions could have an outsize benefit at a population level while reducing logistical hurdles by allotting later administration times to other patients.

These subgroup patterns also raise important biological questions. Our observation that CAR-T cell performance may be more time-sensitive in women is consistent with data emerging with other immunotherapies. This difference warrants further investigation, with particular attention to premenopausal vs postmenopausal women, given that circadian factors interact with sex hormones.^23^

Why anti-CD19 CAR-T cell performance might be particularly time-sensitive is unclear. Since these cells are given to treat lymphomas and leukemias, while anti-BCMA CAR-T cells are used for multiple myeloma, we speculate that tumor-specific factors are responsible. Specifics of CAR-T cell product design and baseline patient risk could also play a role.

We observed that the risk of severe ICANS (and corresponding anakinra use) increased with later CAR-T cell administration times. In contrast, neither severe CRS nor tocilizumab use varied significantly after adjustment for covariates. Several biological mechanisms specific to neurotoxicity could explain this finding. For example, blood-brain barrier permeability varies diurnally in pre-clinical models, which might affect the ability of CAR-T cells to invade the CNS.^29^ Alternatively, circadian rhythms intrinsic to microglia or astrocytes could provoke differing levels of neuroinflammation upon encountering infiltrating CAR T-cells. More research is needed to investigate these possibilities.

This study has several strengths, including two geographically distinct treatment centers demonstrating consistent time-of-day patterns, consideration of both adverse effects and survival outcomes, and rigorous complementary analytical approaches. There are also important limitations. As in any retrospective study, the possibility of unmeasured confounding, including confounding by indication, can never be fully excluded.

Nonetheless, patients receiving late-day CAR T administrations had similar performance status, comorbidity burden, and illness severity compared to patients receiving earlier infusions, and we explicitly adjusted for observable pre-infusion differences (e.g., time in hospital). E-values indicate that substantial unmeasured confounding would be required to nullify our findings.^22^ The reproducibility of these temporal patterns at geographically disparate sites further argues against residual confounding.

While our data estimate associations between time-of-day and CAR-T cell performance at a population level, it cannot estimate risks for individual patients. This is because biological rhythms in any individual can vary with lifestyle factors such as chronotype (e.g., habitual early versus late risers) and night shift work. A precision medicine approach will be needed to address this issue, leveraging actigraphy or other patient-specific data to estimate optimal treatment times.

While our sample size limited the ability to perform more complex analyses with robust statistical power, our two-center cohort is sizable for a study of this relatively new therapy. Although we included a diverse set of clinically important outcomes, we could not evaluate long-term, rare, or emerging negative CAR-T cell outcomes (e.g., CAR-positive peripheral T-cell lymphomas).^30^

Finally, although observational results alone cannot prove causality, pre-clinical evidence supports circadian clock involvement, and the observed morning advantage in CAR-T cell outcomes aligns with predictions based on animal models.^11^ More broadly, the convergence of multiple unrelated immunotherapies – spanning vaccines, checkpoint inhibitors, HSCT, and now CAR-T cells – onto time-of-day patterns in which mornings are clinically optimal further suggests circadian biology may play a fundamental role. In conclusion, our analyses identify time of day as a compelling and easily modifiable candidate factor for optimizing CAR-T cell clinical performance that should be evaluated in future prospective trials.

## Data Availability

All data produced in the present study are available upon reasonable request to the authors

## Contributors

PGL, CAM, and JAH conceptualized the study. EG, MB, PK, BPE, ZL, and BHL extracted the data. PGL, JA H, CH, NS, GH, JH, CAM, and HM performed analyses and interpreted the data. PGL, CAM, HM, and JAH composed the manuscript. All the authors reviewed and edited the manuscript.

## Acknowledgements

We thank Robyn Puro and Carrie Gierasch for their editorial input. This work was supported by NIH K08CA270383 (PGL), NIH R01HL172823 (JAH), NIH R01HL173976 (JAH), and by the Department of Internal Medicine, Washington University School of Medicine (CAM).

## Data Sharing

Requests for de-identified data should be sent to the corresponding authors.

