## Supplemental Appendix for "Diurnal rhythms in chimeric antigen receptor T cell performance: an observational study of 670 patients"

**Contents**

1. STROBE Checklist.
2. eTable 1. Data preparation and missingness handling.
3. eTable 2. Characteristics of patients receiving CAR T-cell infusions before and after 15:00.
4. eTable 3. Adjusted survival outcomes, with exposure considered as time of day as well as time since sunrise.
5. eTable 4. Unadjusted and adjusted odds for secondary (complication) outcomes, with exposure considered as time of day as well as time since sunrise.
6. eTable 5. Laboratory values
7. eFigure 1. Study inclusion details.
8. eFigure 2. Sunrise-adjusted CAR T-cell treatment times vary and are associated with survival
9. eFigure 3. Association between CAR-T cell infusion times and event-free survival.
10. eFigure 4. Association between CAR-T cell infusion times and complications.
11. eFigure 5. Absolute lymphocyte count recovery did not substantially differ by CAR T-cell infusion time of day.
12. eFigure 6. Results of subgroup analyses by hospital and CAR T-cell infusion location.
13. References.

**STROBE Checklist**

| Item | # | Recommendation | Page # | Relevant manuscript text |
|---|---|---|---|---|
| Title | 1A | Indicate the study's design with a commonly used term in the title or the abstract | 1 | Observational cohort study |
| Abstract | 1B | Provide in the abstract an informative and balanced summary of what was done and what was found | 3 | Abstract page |
| Background | 2 | Explain the scientific background and rationale for the investigation being reported | 4 | Introduction section |
| Objectives | 3 | State specific objectives, including any prespecified hypotheses | 4 | …hypothesizing that earlier administration times would correlate with better clinical outcomes and fewer or less severe complications |
| ***Methods*** | | | | |
| Study Design | 4 | Present key elements of study design early in the paper | 4 | Retrospective cohort study |
| Setting | 5 | Describe the setting, locations, and relevant dates, including periods of recruitment, exposure, follow-up, and data collection | 5 | Hospitals, dates, etc. |
| Participants | 6 | Give the eligibility criteria, and the sources and methods of selection of participants. Describe methods of follow-up | 5 | Inclusion/exclusion criteria |
| Variables | 7 | Clearly define all outcomes, exposures, predictors, potential confounders, and effect modifiers. Give diagnostic criteria, if applicable | 6, eTable 1 | All variables described |
| Data Sources | 8 | For each variable of interest, give sources of data and details of methods of assessment (measurement). Describe comparability of assessment methods if there is more than one group | 6 | Data sources, follow-up |
| Bias | 9 | Describe any efforts to address potential sources of bias | 6-7 | Model adjustment |
| Study size | 10 | Explain how the study size was arrived at | 5 | Convenience sample |
| Quantitative variables | 11 | Explain how quantitative variables were handled in the analyses. If applicable, describe which groupings were chosen and why | eTable 1 | Full details in table |
| Statistical Methods | 12A | Describe all statistical methods, including those used to control for confounding | 6-7 | Models, comparisons |
| | 12B | Describe any methods used to examine subgroups and interactions | 7 | Subgroup analyses |
| | 12C | Explain how missing data were addressed | eTable 1 | Imputation strategies |
| | 12D | If applicable, explain how loss to follow-up was addressed | 5 | Follow-up section |
| ***Results*** | | | | |
| Participants | 13 | Report numbers of individuals at each stage of study—eg numbers potentially eligible, examined for eligibility, confirmed eligible, included in the study, completing follow-up, and analysed | 8, eFigure 1 | All details reported |
| Descriptive Data | 14 | Give characteristics of study participants (eg demographic, clinical, social) and information on exposures and potential confounders | 8, Table 1, eTable 1 | |

| | | | | |
|---|---|---|---|---|
| Outcome Data | 15 | Report numbers of outcome events or summary measures over time | 8 | All rates reported |
| Main Results | 16 | Give unadjusted estimates and, if applicable, confounder-adjusted estimates and their precision (eg, 95% confidence interval). Make clear which confounders were adjusted for and why they were included | 8-9 | Model results give categories of confounders, rationale, and results with 95% CI |
| Other Analyses | 17 | Report other analyses done | 9-10 | Subgroup, lab analyses |
| **Discussion** | | | | |
| Key results | 18 | Summarise key results with reference to study objectives | 10 | First paragraph |
| Limitations | 19 | Discuss limitations of the study, taking into account sources of potential bias or imprecision. Discuss both direction and magnitude of any potential bias | 12 | Limitations paragraph |
| Interpretation | 20 | Give a cautious overall interpretation of results considering objectives, limitations, multiplicity of analyses, results from similar studies, and other relevant evidence | 10-12 | Full Discussion |
| Generalizability | 21 | Discuss the generalisability (external validity) of the study results | 10-11 | Discussion of other circadian patterns in immunotherapy, discussion of population-level patterns vs individual-level predictions |
| Funding | 22 | | 1 | Disclosed on title page |

**eTable 1. Data handling, including missingness, for covariates in multivariable models.**

| Variable | Modeling Strategy | Encounters with missing values, n (%) | Value imputed |
|---|---|---|---|
| Age | Continuous | 0 | NA |
| Sex | Dichotomous: Male, Female | 0 | NA |
| Race/Ethnicity | Dichotomous: NonHispanic White, All Others | 23 | Modal value by hospital |
| Cancer Diagnosis | Categorical: Multiple Myeloma, Acute Leukemia, Lymphoma | 0 | NA |
| Lymphodepletion Regimen | Dichotomous: Fludarabine/Cyclophosphamide, Other | 0 | NA |
| Performance status | Continuous | 9 | Median ECOG by hospital, malignancy, and CAR T-cell product |
| Van Walraven score | Continuous | 71 | Median value by hospital |
| SAPS-2 | Continuous, with age component excluded in models | 0 | NA |
| CAR T-cell dose | Continuous standardized by malignancy, product, and body weight | 186 | Median dose by hospital, malignancy, and CAR T-product |
| Season | Categorical: Spring, Summer, Fall, Winter | 0 | NA |
| Time period | Categorical: 2018-2019, 2020-2021, 2022-2024 | 0 | NA |
| Days in hospital prior to CAR T-cell administration | Continuous, defaulting to 0 for outpatient administrations | 0 | NA |
| ECOG, Eastern Cooperative Oncology Group; CAR (chimeric antigen receptor) T-cell; SAPS-2 (Simplified Acute Physiology Score) | | | |

**eTable 2. Characteristics of patients with early (before 15:00) and late (after 15:00) CAR T-cell infusions.**

| Characteristic | 08:00-14:59, N = 635 | 15:00-18:00, N = 35 |
|---|---|---|
| Hospital, N (%) | | |
| WU | 345 (54%) | 18 (51%) |
| OHSU | 290 (46%) | 17 (49%) |
| Study Period, n (%) | | |
| 2018-2019 | 95 (15%) | 4 (11%) |
| 2020-2022 | 267 (42%) | 14 (40%) |
| 2023-2024 | 273 (43%) | 17 (49%) |
| Season, n (%) | | |
| Fall | 158 (25%) | 4 (11%) |
| Spring | 168 (26%) | 13 (37%) |
| Summer | 164 (26%) | 9 (26%) |
| Winter | 145 (23%) | 9 (26%) |
| Age, median (IQR) | 65 (57, 72) | 60 (39, 72) |
| Female, n (%) | 214 (34%) | 8 (23%) |
| Malignancy, n (%) | | |
| DLBCL | 392 (62%) | 18 (51%) |
| Follicular Lymphoma | 21 (3·3%) | 2 (5·7%) |
| Leukemia | 31 (4·9%) | 4 (11%) |
| MCL | 54 (8·5%) | 2 (5·7%) |
| Multiple Myeloma | 137 (22%) | 9 (26%) |
| CAR T-cell Product, n (%) | | |
| Axi-cel | 241 (38%) | 9 (26%) |
| Brexu-cel | 70 (11%) | 3 (8·6%) |
| Cilta-cel | 73 (11%) | 3 (8·6%) |
| Ide-cel | 64 (10%) | 6 (17%) |
| Liso-cel | 73 (11%) | 6 (17%) |
| Tisa-cel | 114 (18%) | 8 (23%) |
| Flu/Cy Conditioning, n (%) | 527 (83%) | 28 (80%) |
| Hospital Days Before Infusion, median (IQR) | 1 (1, 1) | 1 (1, 5) |
| ECOG Performance Status, median (IQR) | 1 (0, 1) | 1 (1, 1) |
| Van Walraven Score, median (IQR) | 9 (6, 15) | 12 (9, 19) |
| SAPS-2, median (IQR) | 28 (24, 32) | 29 (23, 34) |

CAR T-cell, chimeric antigen receptor T-cell; WU, Washington University; OHSU, Oregon Health & Science University; IQR, interquartile range; DLBCL, diffuse large B-cell lymphoma; Flu, fludarabine; Cy, cyclophosphamide; ECOG, Eastern Cooperative Oncology Group; SAPS-2, Simplified Acute Physiology Score.

**eTable 3. Adjusted survival outcomes, where the exposure is a one-hour shift in CAR-T cell infusion times (in units of clock time or indexed to the local time of sunrise).**

| | Exposure = One Hour Shift (Clock Time) | | | | | Exposure = One Hour Shift (Adjusted to Local Sunrise) | | | | |
|---|---|---|---|---|---|---|---|---|---|---|
| **Outcome** | **aOR** | **Lower CI** | **Upper CI** | **p-value** | **E-value** | **aOR** | **Lower CI** | **Upper CI** | **p-value** | **E-value** |
| ***Adjusted mixed-effects multivariable logistic regression models*** | | | | | | | | | | |
| 90-day OS | 0·783 | 0·663 | 0·924 | 0·004 | 1·51 | 0·788 | 0·670 | 0·926 | 0·004 | 1·50 |
| 90-day EFS | 0·902 | 0·763 | 1·07 | 0·225 | NA | 0·884 | 0·764 | 1·020 | 0·096 | NA |
| 365-day OS | 0·835 | 0·735 | 0·948 | 0·005 | 1·42 | 0·826 | 0·730 | 0·935 | 0·003 | 1·433 |
| 365-day EFS | 0·874 | 0·744 | 1·030 | 0·102 | NA | 0·867 | 0·748 | 1·000 | 0·057 | NA |
| ***Adjusted linear pseudo-value time-to-event models*** | | | | | | | | | | |
| | **RMST Difference** | **Lower CI** | **Upper CI** | **p-value** | **E-value** | **RMST Difference** | **Lower CI** | **Upper CI** | **p-value** | **E-value** |
| 2-year overall RMST | -21·8 days | -36·4 days | -8·03 days | NA* | NA* | -20·8 days | -34·6 days | -7·13 days | NA* | NA* |
| 2-year event-free RMST | -18·0 days | -34·2 days | -2·21 days | NA* | NA* | -17·2 days | -32·3 days | -1·53 days | NA* | NA* |

RMST, restricted mean survival time; CI, confidence interval; aOR, adjusted odds ratio; OS, overall survival; EFS, event-free survival
*Adjusted RMST models were estimated using pseudo-values within a linear mixed-effects framework with robust standard errors. As this approach does not operate on the log-odds scale and is not likelihood-based, standard p-value-based hypothesis testing and E-values are not applicable; results are presented as absolute survival differences to emphasize clinical relevance. Negative RMST differences indicate lower RMST with later-day CAR T-cell administrations.

**eTable 4. Adjusted secondary (complication) outcomes, where the exposure is a one-hour shift in CAR-T cell infusion times (in units of clock time or indexed to the local time of sunrise).**

| | Exposure = One Hour Shift (Clock Time) | | | | | Exposure = One Hour Shift (Adjusted to Local Sunrise) | | | | |
|---|---|---|---|---|---|---|---|---|---|---|
| **Outcome** | **aOR** | **Lower CI** | **Upper CI** | **p-value** | **E-value** | **aOR** | **Lower CI** | **Upper CI** | **p-value** | **E-value** |
| ***Adjusted mixed-effects multivariable logistic regression models*** | | | | | | | | | | |
| ICU Admission | 1·03 | 0·834 | 1·27 | 0·793 | NA | 1·04 | 0·865 | 1·25 | 0·667 | NA |
| Mechanical Ventilation | 1·29 | 1·03 | 1·63 | 0·0289 | 1·53 | 1·28 | 1·02 | 1·6 | 0·0348 | 1·51 |
| Vasopressors | 1·13 | 0·937 | 1·37 | 0·197 | NA | 1·12 | 0·931 | 1·34 | 0·232 | NA |
| Infection | 1·14 | 0·987 | 1·31 | 0·0747 | NA | 1·15 | 0·998 | 1·31 | 0·0531 | NA |
| CRS 3/4 | 0·956 | 0·744 | 1·23 | 0·728 | NA | 0·974 | 0·771 | 1·23 | 0·824 | NA |
| ICANS 3/4 | 1·17 | 1·01 | 1·34 | 0·0309 | 1·359 | 1·17 | 1·02 | 1·33 | 0·0269 | 1·37 |
| Tocilizumab | 1·02 | 0·906 | 1·14 | 0·766 | NA | 1·01 | 0·898 | 1·13 | 0·925 | NA |
| Anakinra | 1·26 | 1·07 | 1·49 | 0·00645 | 1·5 | 1·26 | 1·07 | 1·48 | 0·00505 | 1·5 |
| aOR, adjusted odds ratio; CI, confidence interval; CRS, cytokine release syndrome; ICANS, immune cell-associated neurotoxicity syndrome | | | | | | | | | | |

**eTable 5: Maximal values for selected inflammatory serologies after early (before 15:00) and late (after 15:00) CAR-T Cell infusions.**

| Test | Early CAR-T cell infusion, median (IQR)[1] | Late CAR-T cell infusion, median (IQR)[2] |
|---|---|---|
| CRP | 112 (31-198) | 126 (53-259) |
| Ferritin | 1005 (400-2759) | 1759 (400-4125) |
| LDH | 305 (250-412) | 333 (250-673) |
| IFN-$\gamma$ | 4.6 (4.2-17) | 4.7 (4.2-22) |
| IL-10 | 37 (17-128) | 36 (13-267) |
| IL-13 | 1.7 (1.7-4.2) | 2 (1.7-4.4) |
| IL-17 | 1.4 (1.4-1.5) | 1.4 (1.4-2.2) |
| IL-2 | 5.3 (2.1-16) | 5.2 (2.1-16) |
| IL-4 | 2.2 (2.2-2.2) | 2.2 (2.2-2.2) |
| IL-5 | 11 (3.2-39) | 10 (6.4-125) |
| IL-6 | 54 (11-341) | 98 (4-711) |
| IL-8 | 3 (3-3) | 3 (3-3) |
| TNF-$\alpha$ | 2.6 (1.7-4.9) | 2.3 (1.7-7.1) |

[1]n=627 for early CAR-T cell infusions and n=34 for late CAR-T cell infusions.

[2]n=154 for early CAR-T cell infusions and n=13 for late CAR-T cell infusions.

**eFigure 1. Flow diagram.**

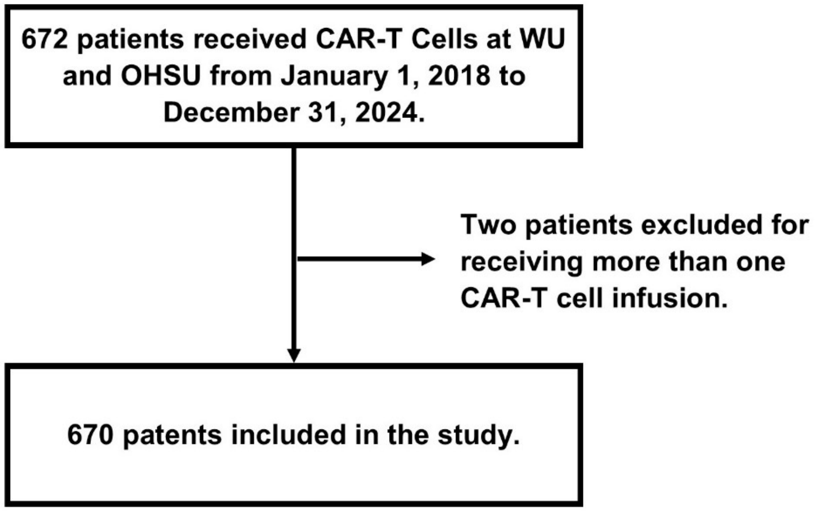

**eFigure 2. Sunrise-adjusted CAR T-cell treatment times vary and are associated with survival. A** depicts the distribution of CAR T-cell infusion times (since sunrise) at WU (red bars) and OHSU (blue bars). **B** is a multivariable hierarchical logistic regression analysis depicting adjusted 90-day (black line) and 365-day (green line) overall survival as a function of CAR T-cell infusion time since sunrise. The shaded area represents 95%-CIs.

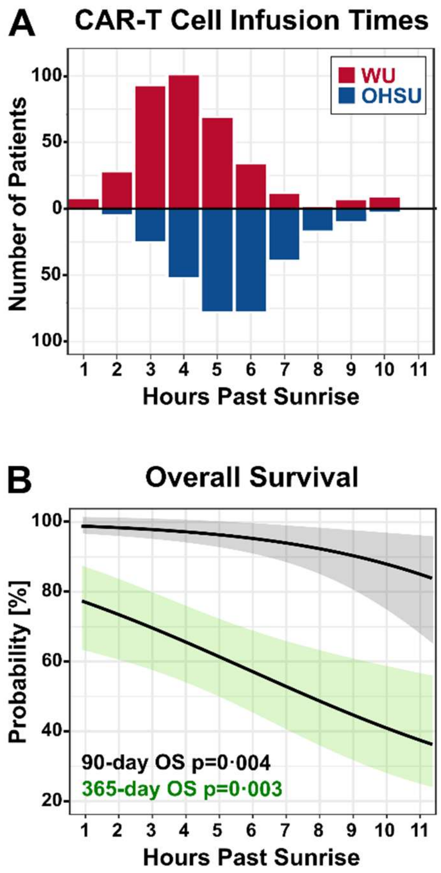

**eFigure 3. Association between CAR-T cell infusion times and event-free survival.** The panels depict multivariate regression analyses correlating 90-day EFS (top row) and 365-day EFS (bottom row) with CAR-T infusion time, expressed as hours of the clock (left column) or hours since local sunrise (right column). Shaded areas represent 95%-CIs. Wald test calculated p-values are depicted.

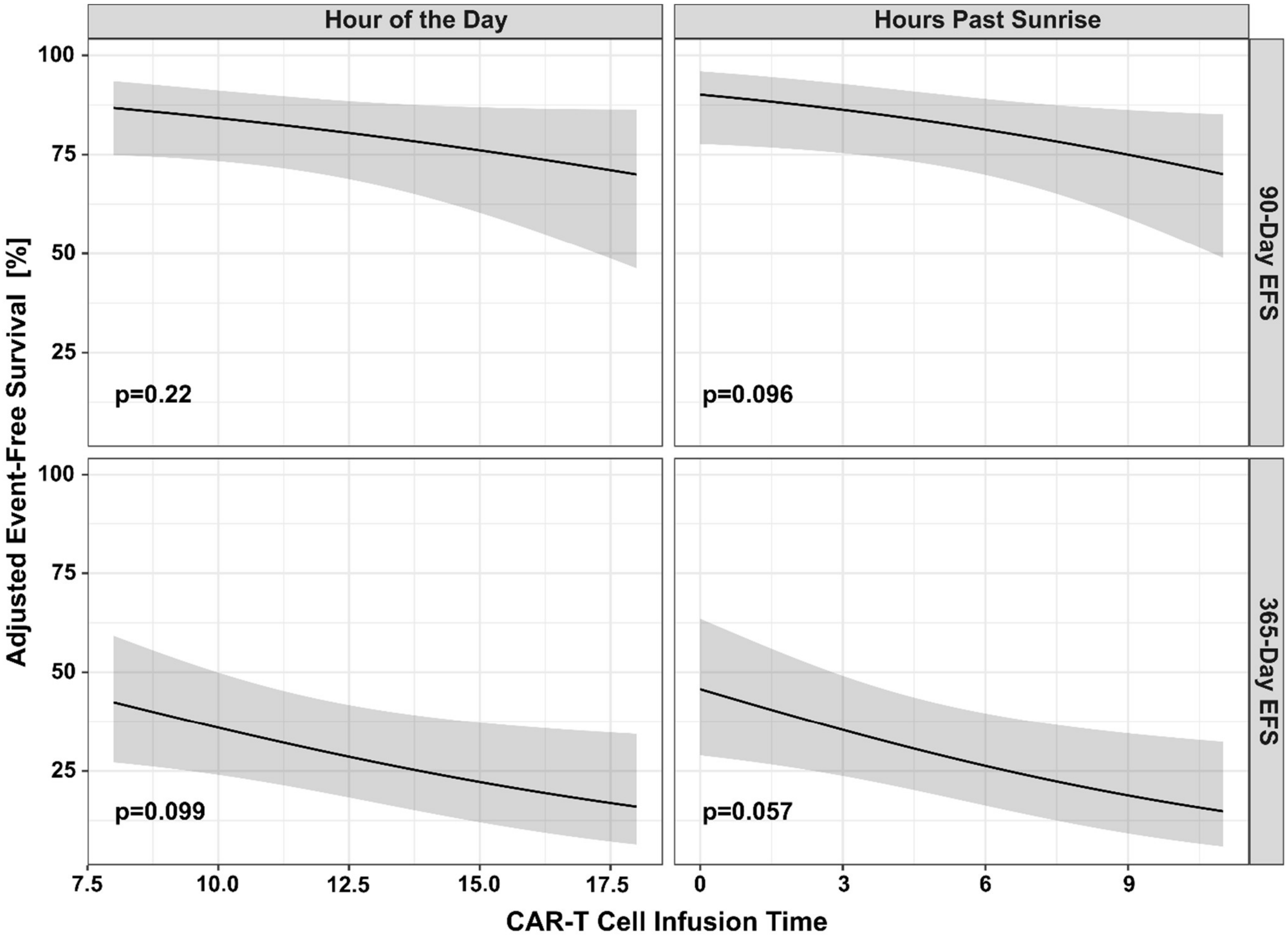

**eFigure 4. Association between CAR-T cell infusion times and complications**. The panels depict multivariate regression analyses correlating the probability of selected complications with CAR-T infusion time, expressed as hours of the clock (left column) or hours since local sunrise (right column). Each row represents a different complication as labeled to the right. Shaded areas represent 95%-CIs. Wald test calculated p-values are depicted. p-values are derived from approximate Wald tests based on the estimated covariance of the spline coefficients.

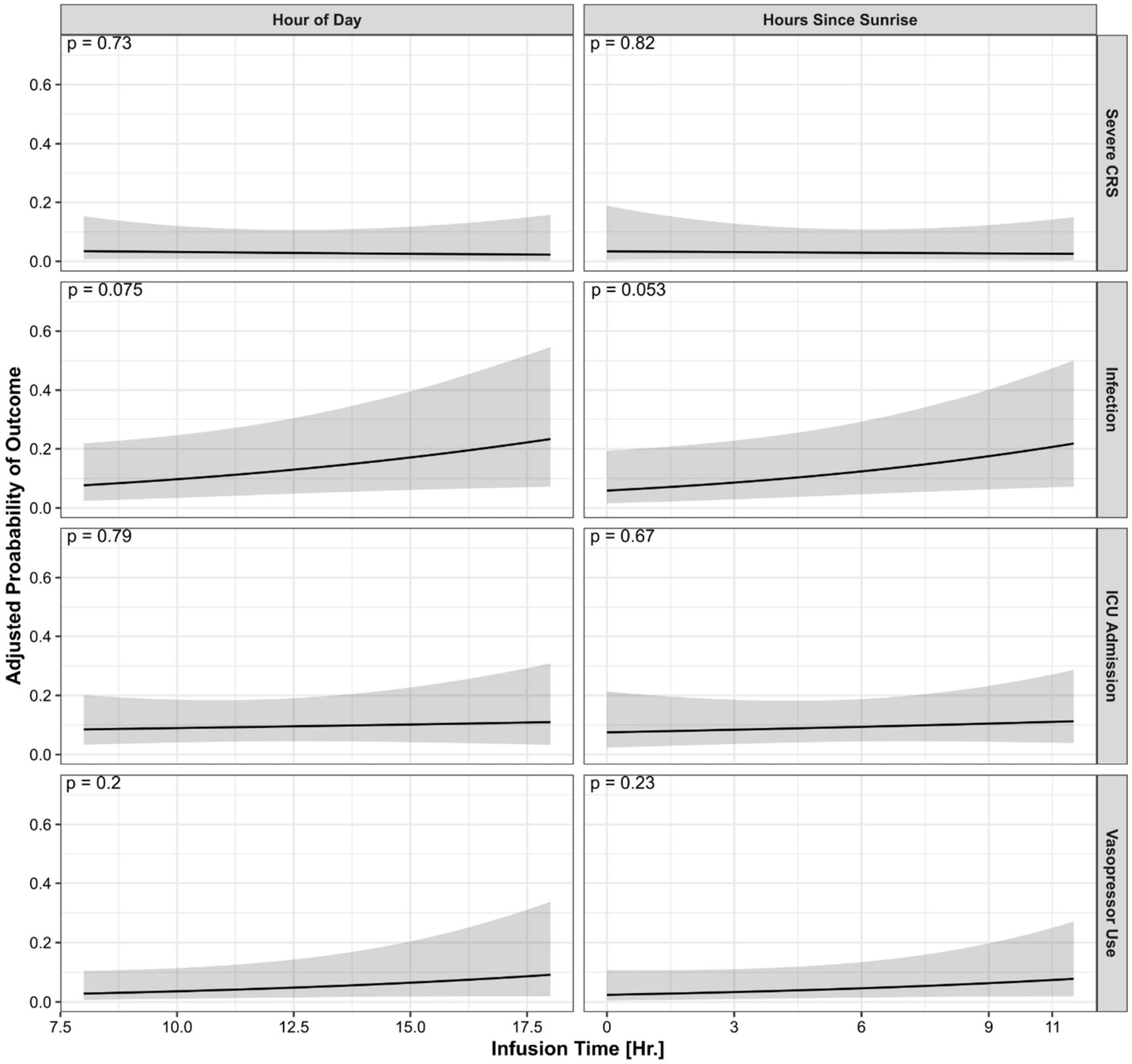

**eFigure 5. Absolute lymphocyte count recovery did not substantially differ by CAR T-cell infusion time of day. The x-axis represents time, in days, since CAR T-cell infusion.** The y-axis represents the absolute lymphocyte count. Orange indicates patients infused before 15:00 and purple indicates those infused at or after 15:00. Each value is one scattered point. Boxes represent the interquartile range (IQR) with the horizontal line indicating the median; whiskers extend to 1·5 × IQR.

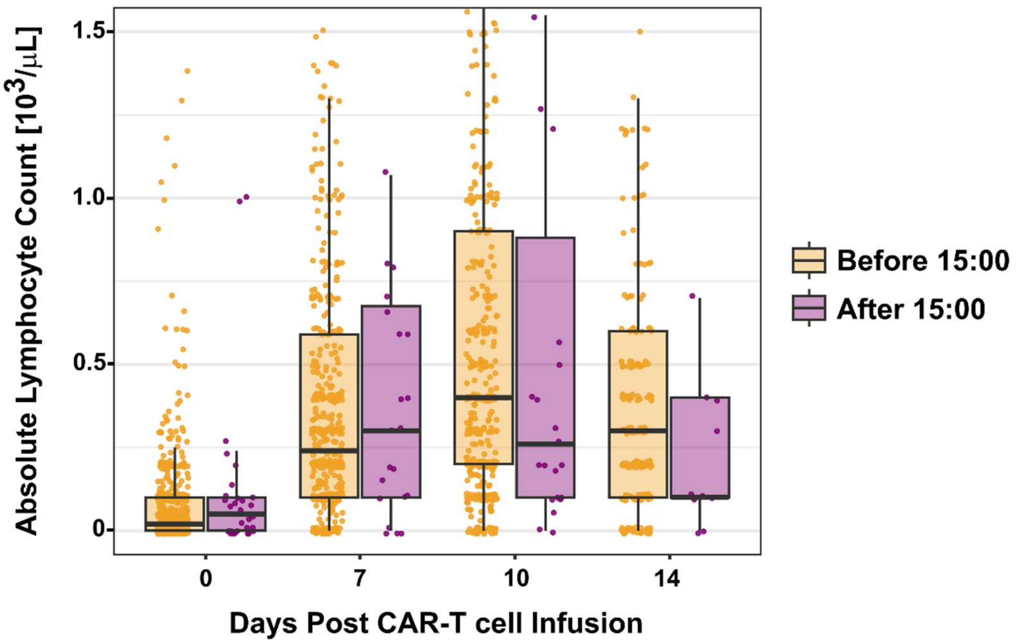

**eFigure 6. Results of subgroup analyses by hospital and CAR T-cell infusion location**. Within each panel (left = hospital [red = WU, blue = OHSU], right = inpatient [pink] vs outpatient [purple]), the x-axis represents hourly adjusted odds ratios (aORs) for landmarked survival outcomes (y-axis). aOR estimates are represented by points, with 95% CI as horizontal lines. Values to the left of the vertical dashed line indicate that the outcome would be more likely with earlier CAR T-cell infusions, while values to the right of the line suggest the outcome would be more likely with later infusions.

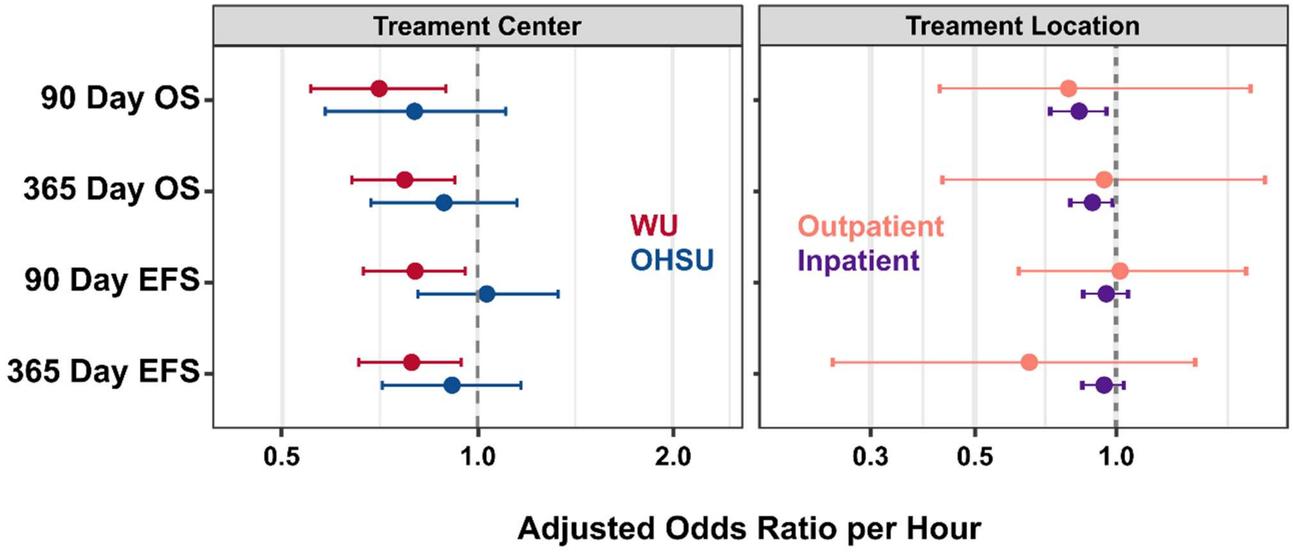
